# Benchmarking open-source automated thigh muscle MRI segmentation algorithms

**DOI:** 10.64898/2026.09.16.26363275

**Authors:** Candace Makeda Moore, Morris Alper

**Affiliations:** Adekam, Zaandam, North Holland, Netherlands; Department of Computer Science, University of Miami, Miami, Florida, USA

## Abstract

Automatic segmentation of medical images could accelerate clinical and research workflows, yet existing tools are rarely compared independently on shared benchmarks, making it difficult to assess genuine progress. This is particularly consequential for limb muscle segmentation in magnetic resonance imaging (MRI), which underpins volumetric analysis, radiomics, and fat fraction quantification used as biomarkers in neuromuscular disease – applications where segmentation errors can directly affect interpretation. Despite the availability of multiple open-source segmentation tools spanning diverse architectures, no independent head-to-head comparison has appeared in the literature. We address this gap by evaluating eight tools, six of which run independently, and the other two of which are meant to be auxiliary algorithms once muscles have been somehow initially labeled. We evaluated these tools on three-dimensional MRI volumes across three base datasets and then later against derived datasets, assessing both quantitative metrics and qualitative usability. Our results reveal substantial performance variation across methods and datasets, with many methods showing reduced accuracy on pathological cases. Domain-specific models trained on large datasets consistently outperformed foundation models and newer general-purpose architectures. Our results reveal practical trade-offs between accuracy, generalizability, and ease of use that are critical for clinical adoption, while raising concerns for deployment on underrepresented populations and pathologies.

## Introduction

Medical segmentation—the delineation of anatomical structures in medical images—is foundational to tasks ranging from radiotherapy planning [1, 2] to surgical navigation [3] and longitudinal assessment of tissue changes. In limb muscle MRI, segmentation enables volumetric analysis, radiomics, and per-muscle fat fraction quantification, which serve as biomarkers for neuromuscular and musculoskeletal diseases [4]. Manual segmentation remains common but is time-consuming, operator-dependent, and poorly reproducible [4, 5], creating a bottleneck for large-scale studies. This is especially acute for rare diseases, where manual segmentation remains the norm due to limited and unrepresentative training data [6, 7]. Automatic segmentation could in principle address these limitations, but only if it is accurate enough to be trusted without extensive manual correction, while inaccurate automation risks introducing systematic errors while creating a false sense of reliability.

A variety of open-source tools for limb muscle segmentation have become available in recent years, spanning architectures from U-Nets trained on large curated datasets [8] to alternative architectures such as HRNet variants [9] and many others, though recent evidence suggests U-Nets generally remain competitive for 3D medical image segmentation [10]. While the search for better algorithms appropriate for various tasks continues, some newer algorithms have not yet been developed as open-source tools for thigh segmentation. While diffusion models are considered an important novel paradigm for segmentation, and we found medical imaging diffusion models existed [11], [12] , they had not been applied to thigh MRI in an open framework as of yet. Few-shot and other low-data approaches have also been explored but remain limited in practical applicability for this domain [13–15]. As machine learning has moved towards using foundation models [16]—general-purpose neural networks pretrained on diverse, web-scale data and applied to a wide range of downstream tasks—there has been emerging interest in their application to medical imaging. Segment Anything (SAM) [17] in particular has attracted attention as a potentially general-purpose segmentation solution. However, the effectiveness of foundation models on specialized tasks with limited data remains uncertain, even when fine-tuned on domain-specific data [18]. A practical limitation is that SAM operates on 2D slices, requiring volumetric images to be processed slice-by-slice and re-concatenated. Early evaluations found that SAM underperformed domain-specific models on medical images [19], and subsequent variants including short-long memory SAM [20] have shown only incremental improvements. Comparisons of SAM variants with U-Nets on neuromuscular disease data have found mixed results [21]. Thus the question of whether a better architecture than U-Net has emerged is open. Existing surveys either predate key foundation models [22] or compare methods only qualitatively without quantitative benchmarking [23].

Unfortunately a fundamental difficulty in assessing progress is that many existing medical segmentation tools were built for different use cases; thus differing evaluation data and metrics prevent direct comparison of results. Further complicating any attempt at evaluation is that most evaluation surveys look at very different anatomy and pathology e.g. brain tumors, and therefore results would not be expected to be indicative of performance unless the models were retrained on limb data. Ultimately, clinicians and researchers selecting a segmentation tool for limb muscle MRI have no independent basis for comparison.

We address this gap with a systematic, independent benchmark of eight open-source segmentation tools evaluated across multiple datasets, including both healthy and pathological cases. We assess performance via quantitative and qualitative evaluation of segmentation outputs and practical usability. Our aim is to provide the independent comparison needed to guide tool selection and to identify where current methods fall short, particularly for pathological cases, out-of-distribution data, and the practical usability considerations that determine whether a tool can realistically be adopted by clinical researchers without software engineering expertise. We chose the thigh as the anatomy on which to assess tools because lower-limb muscle involvement is common and clinically relevant across many neuromuscular disorders, and the thigh has a higher level of anatomical complexity than the calf ( which has eleven muscles as opposed to the thirteen of the thigh). We find that domain-specific U-Net architectures — particularly MuscleMap — consistently outperform foundation-model-based approaches, and that augmenting existing tools with SAM variants often degrades rather than improves segmentation quality. Close qualitative analysis reveals that the best performing evaluated tools show reduced performance on pathological data, underscoring the need for careful, dataset-specific validation before clinical or research adoption.

## Materials and methods

We benchmark eight open-source segmentation tools against expert ground-truth annotations across multiple MRI datasets. We include all open-source tools we identified that perform individual muscle segmentation of the thigh; we excluded tools that segment only broader compartments rather than individual muscles (e.g., TotalSegmentator MRI), as well as any closed-source or proprietary tools. We evaluate open-source tools because they are available to all research groups, including those in hospitals in lower-income countries or otherwise lower-resourced groups, and allow full exploration of code and models. No spatial registrations were applied between ground-truths and predicted segmentations.

### Datasets

We began testing on three base datasets: MyoSegmenTUM [24], AIPS [25] and Sheffield [26]. All three datasets had already been de-identified prior to release under each source study’s own ethics approval; no additional identifiable information was collected or handled for this benchmark. In a further round of testing we used datasets derived from MyoSegmenTUM created to test robustness to pathological imaging and out-of-distribution data. The original base datasets are summarized in Table 1:

**Table 1.**
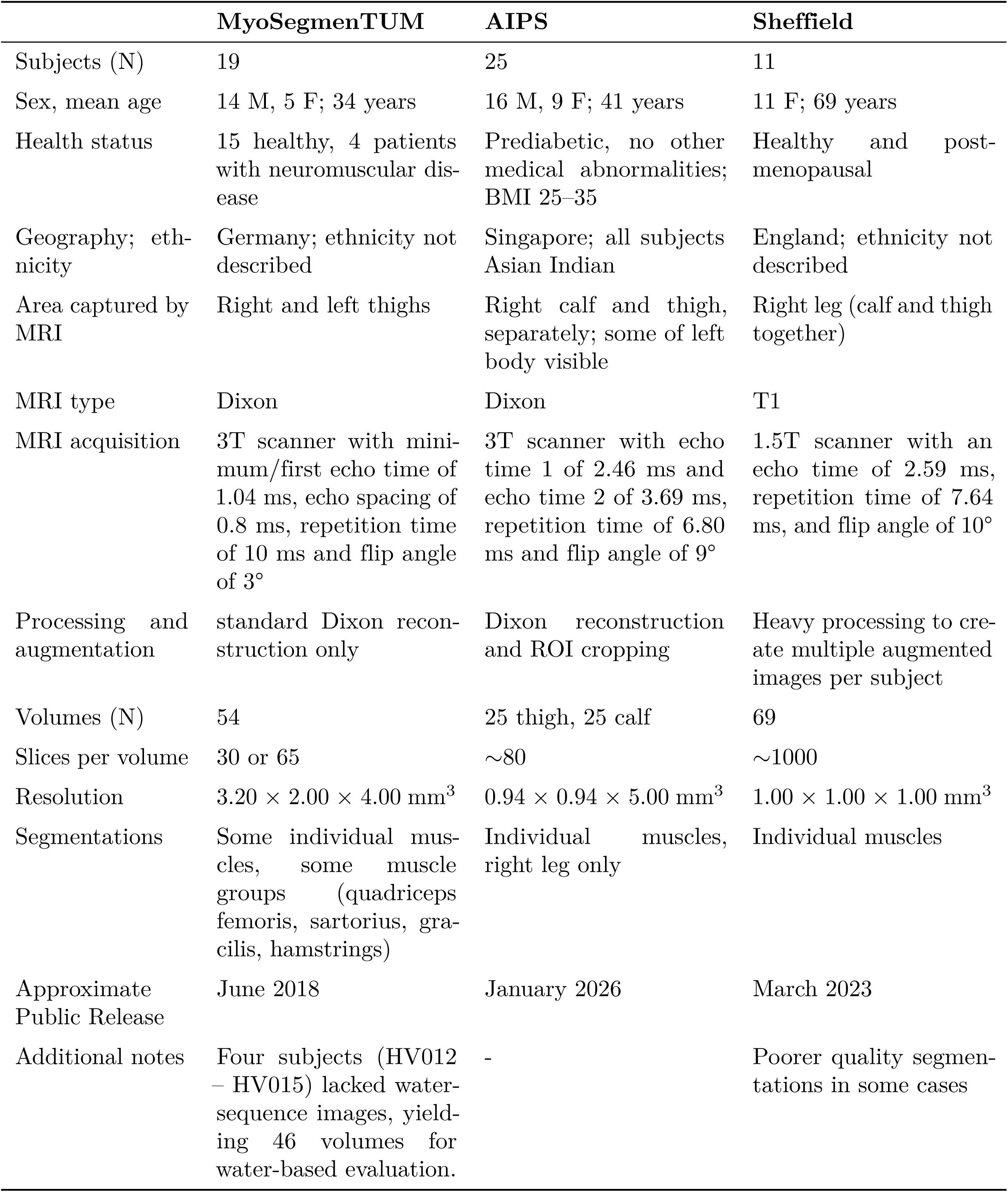
Overview of three base MRI datasets used in this benchmark.

We observed some issues with the segmentation of the Sheffield dataset [26] [27] [28], and have included images which illustrate the problem: (Figures 1 and 2)

**Fig 1.**
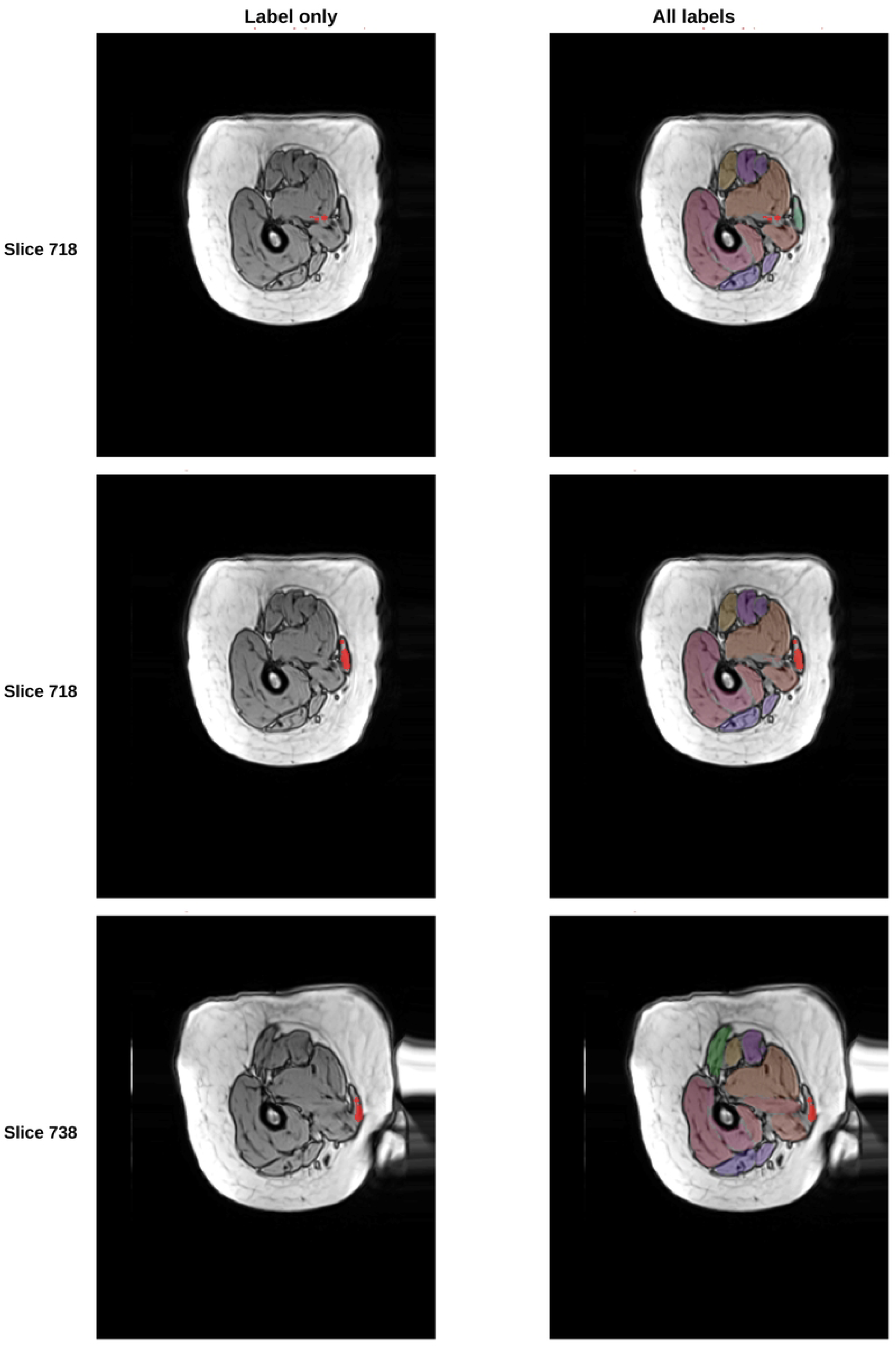
Images from Sheffield dataset sample Aug-2 demonstrating poor labeling over the course of several muscles. The labeled muscles are shown in red. The left column shows the individual muscle label, while the right column shows all labels. The top row shows adductor brevis at slice 718, which is poorly labeled. The middle row shows gracilis at slice 718 with questionable labeling, while the bottom row shows gracilis at slice 738 with incomplete segmentation. The poor labeling of both muscles was present across multiple slices.

**Fig 2.**
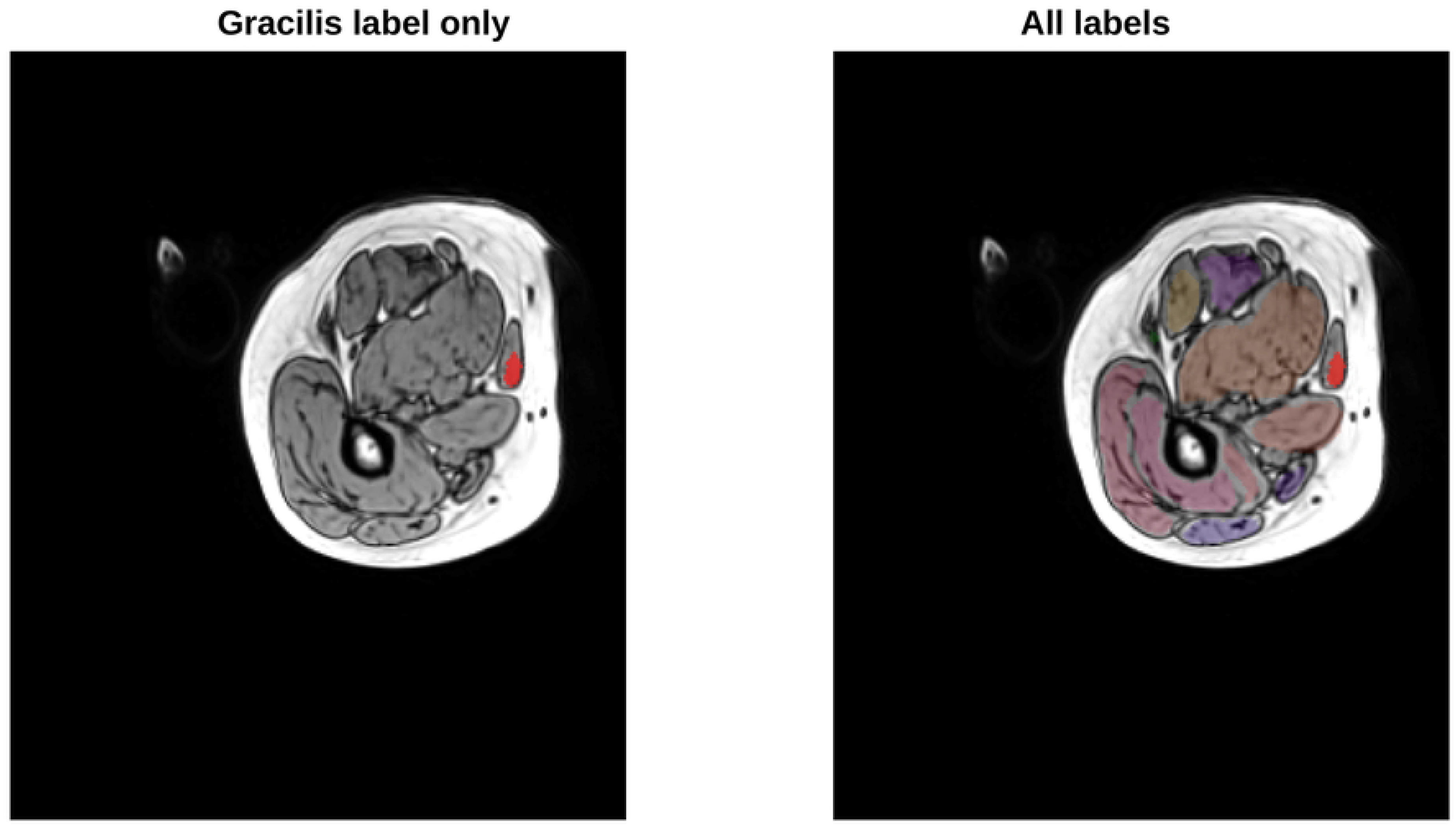
Images from Sheffield dataset, Aug-8, slice 644, showing gracilis segmentation in red which is incomplete. Left image with only gracilis label, right image with all labels. Note adductor magnus segmentation directly below indicated with red arrow/caret goes beyond muscle and therefore incorrectly labels fat as muscle.

We derived two additional datasets from the MyosegmenTUM dataset to test robustness:

### Pathological Subset of MyoSegmenTUM [24]

A subset of MyoSegmenTUM of 12 bilateral thigh MRI volumes with expert muscle segmentations, providing both fat fraction and water sequences from the four subjects (P001–P004) with neuromuscular diseases. The diseases included myotonic dystrophy type 2 (n =2), limb girdle muscular dystrophy 2A (n = 1), and amyotrophic lateral sclerosis (n = 1). These diseases induce predictable patterns of different changes in various muscles-with some muscles relatively spared in terms of pathological processes like fatty infiltration or atrophy, compared to others. However the predicted and real changes vary from patient to patient on a macroscopic, and thus MRI level. Thus it is not possible even on this small subset of pathological imaging to simply use quantitative metrics on the most affected muscles, as they differ per patient. In this context relevant analyses must weigh qualitative trends as overall metrics can obscure poor performance on the muscles most affected by disease.

### Augmented MyoSegmenTUM

A dataset derived from original images of the four diseased subjects, and original images of six healthy subjects. Augmented image pairs were generated from the water and fat-fraction NIfTI volumes of each selected subject using a fixed pipeline on the dissector library [29], applying random augmentations (flips, affine and elastic deformations, intensity augmentations) to water and fat-fraction images, with segmentation masks deformed to preserve alignment. Each image was then checked by hand to make sure it appeared as though it was an image in line with general quality tests performed during typical hospital imaging, and would have been an acceptable image not retaken due to poor quality.

This dataset tests robustness to distribution shift, as the augmented images are unlikely to match any tool’s training data.

We selected MyoSegmenTUM for augmentation due to its clean annotations and presence of pathologies; by contrast, AIPS is a pre-diabetic cohort without confirmed pathological muscle changes, and Sheffield does not contain pathologies and has the ground-truth annotation errors noted above; augmenting either would not test the case we are most interested in—muscles altered by disease. This dataset is therefore designed to test robustness to distribution shift specifically for pathological presentations, complementing the Pathological Subset above with images that are further out-of-distribution from any tool’s training data.

### Evaluation protocol

Predicted segmentations were compared to ground truth using code based on the SimpleITK [30] (v2.5.3) library. Primary metrics included the Dice coefficient, Jaccard index, and Hausdorff distance. We also reported volume similarity, false negative rate, false positive rate, binary cross-entropy, boundary intersection over union, and the inter-slice Dice ratio. The Dice similarity coefficient is a recommended and popular metric [31], so we use it both in its standard form and to assess slice-to-slice variability via the inter-slice Dice ratio. This inter-slice variability was of particular concern for algorithms that segment each 2D slice independently, which risks producing an incoherent 3D volume, despite muscle anatomy naturally tapering or expanding gradually from slice to slice. All qualitative figures in this paper display axial slices only, which is a deliberate choice rather than an oversight: axial is the plane in which radiologists and researchers primarily read thigh muscle MRI. We assess cross-slice (through-plane) consistency quantitatively via the inter-slice Dice ratio rather than through sagittal, coronal, or 3D renderings. It should be noted that only one of the datasets is acquired at an isometric resolution, and resolutions such as those in AIPS (0.94 × 0.94 × 5.00 mm) are not uncommon in neuromuscular datasets and make the axial plane the obvious choice for most interpretations.

As Müller et al. explain in their paper on segmentation metrics [31], each metric captures a different aspect of segmentation quality; thus, performance cannot be reduced to a single metric, or even a composite metric, without losing important information about how an algorithm performs.

All tools were evaluated on the full MyoSegmenTUM dataset. Any variation of SAM requires user input, in the form of points or bounding boxes prompts, and is therefore not truly fully automated. All tools capable of independent operation (requiring only unmarked, unsegmented images as input as opposed to algorithms that need some labeling i.e. SAM variants) were further evaluated on the AIPS and Sheffield, and later augmented and pathological datasets.

Our initial evaluations of SAM had shown that the underlying prompts changed performance significantly even if they were semantically equivalent i.e., different points all within a structure. Nonetheless MedCLIP-SAMv2 was also evaluated on all datasets. This meant that all six independent tools but not pure SAM variations were evaluated on all datasets.

For MedCLIP-SAMv2, which requires text prompts, we used the prompt ‘‘[muscle name] [side] thigh Dixon MRI axial cross section,” following the prompt structure used in the authors’ own released examples [32]. The authors quantify the effect of prompt choice on performance (their Table 3), confirmed by our informal experiments.

For Dafne, we ran the distributed model without its interactive correction workflow, while also verifying that segmentation ran via script exactly matched GUI outputs.

All evaluation code and augmentation pipelines are publicly available [29].

### Algorithms

We experimented with training some frameworks for novel algorithms. Specifically, diffusion models, as recent work suggests diffusion-based approaches can be competitive given sufficient data and compute [33]. Using MedSegDiff [34], [29], we eventually judged that our potential training data was not large enough to create representative examples of performance in this category. MuscleSeg [25], a recently released tool which is fairly novel in its use of ensemble learning , was excluded on two grounds: it does not meet our open-source criterion, as its code repository has no license, and its training data was exclusively the AIPS dataset used here. We eventually selected eight already existing tools for thigh MRI. The eight tools evaluated span U-Net variants, foundation models, a federated learning framework, and a multimodal encoder text-driven vision–language framework that couples a biomedical vision–language model with SAM. They are summarized in Table 2:

**Table 2.**
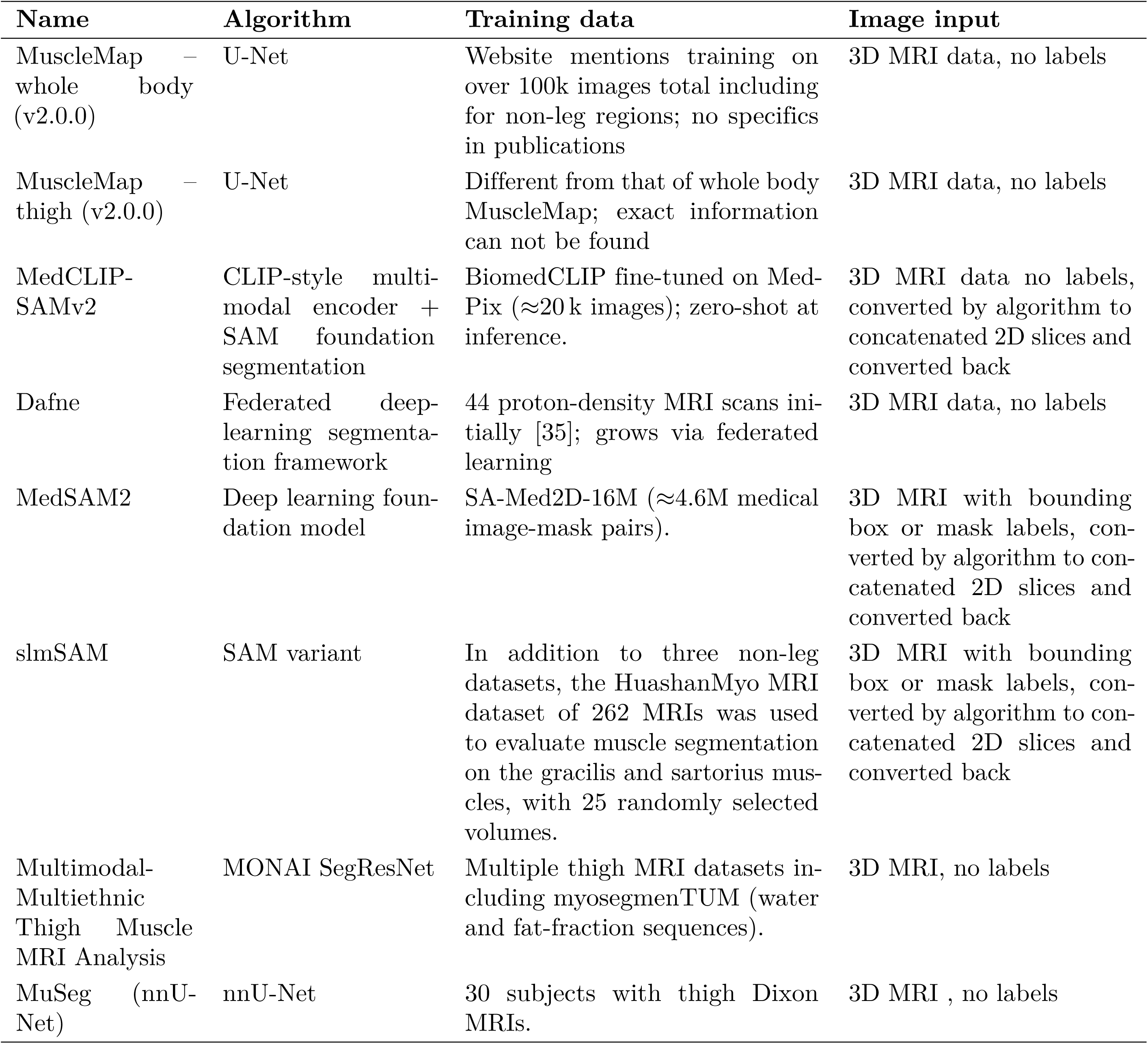
Overview of evaluated thigh muscle segmentation algorithms.

Several tools were tested in multiple configurations. MuscleMap provides both whole-body and thigh-specific models; both were evaluated. MuscleMap whole body is a MONAI (v1.3.2) U-Net with no modifications and num res units (variable) set to 1, whereas the thigh configuration is identical except that num res units is set to 2 (and potential differences in training data to build the model). SAM-based tools (MedSAM2, slmSAM) require point or bounding-box prompts, which we generated automatically from MuscleMap predictions (selected for its low false-positive rate).

MedCLIP-SAMv2 is not a segmentation network in itself but a framework that couples a biomedical vision–language model with SAM [32]. A text prompt is encoded alongside the image by a CLIP-style biomedical encoder, and the resulting cross-modal attribution is converted into a coarse saliency map indicating the region the text refers to. This map is thresholded to produce point or bounding-box prompts, which are then passed to SAM to generate the final mask. Segmentation quality therefore depends jointly on the text prompt, the saliency-to-prompt conversion, and SAM’s response to those prompts.

An important consideration is potential data leakage. As seen in Table 3, at least one tool under consideration (Multimodal-Multiethnic) was trained on data including MyoSegmenTUM. It should therefore be noted that the results of the Multimodal-Multiethnic algorithm on MyoSegmenTUM and datasets derived from it can reflect data leakage because as noted in Table 3, this was a training dataset for that algorithm. For MuscleMap and Dafne, training data descriptions are incomplete, and we cannot confirm whether MyoSegmenTUM or other evaluation datasets were in-distribution.

**Table 3.** In- (✓) and out-of-distribution (*×*) data for algorithms and datasets used for benchmarking. A question mark (?) indicates that this is unknown. For MedCLIP-SAMv2 “*×^∗^*” indicates possible exposure through the BiomedCLIP backbone pretraining corpus (which pre-dates AIPS).

| Name | MyoSegmenTUM | AIPS | Sheffield |
| --- | --- | --- | --- |
| MuscleMap variants | ? | ? | ? |
| MedCLIP-SAMv2 | ×* | × | ×* |
| Dafne | ? | ? | ? |
| MedSAM2 | ? | ? | ? |
| slmSAM | ? | ? | ? |
| Multimodal-Multiethnic | ✓ | × | × |
| MuSeg | × | × | × |

### Artificial Intelligence Tools and Technologies Usage

Generative AI, specifically Claude [36], a large language model, was used to help copy-edit this article. The accompanying software package, Dissector, has some code created with the aid of generative AI. The specific tool used was Claude Code [37]. All code on the main branch and in releases is human author written and/or reviewed.

## Results

We proceed to visualize the main metrics per dataset and independent algorithm in Figure 3, with all quantitative results including additional metrics provided as tables in the supplementary material.

**Fig 3.**
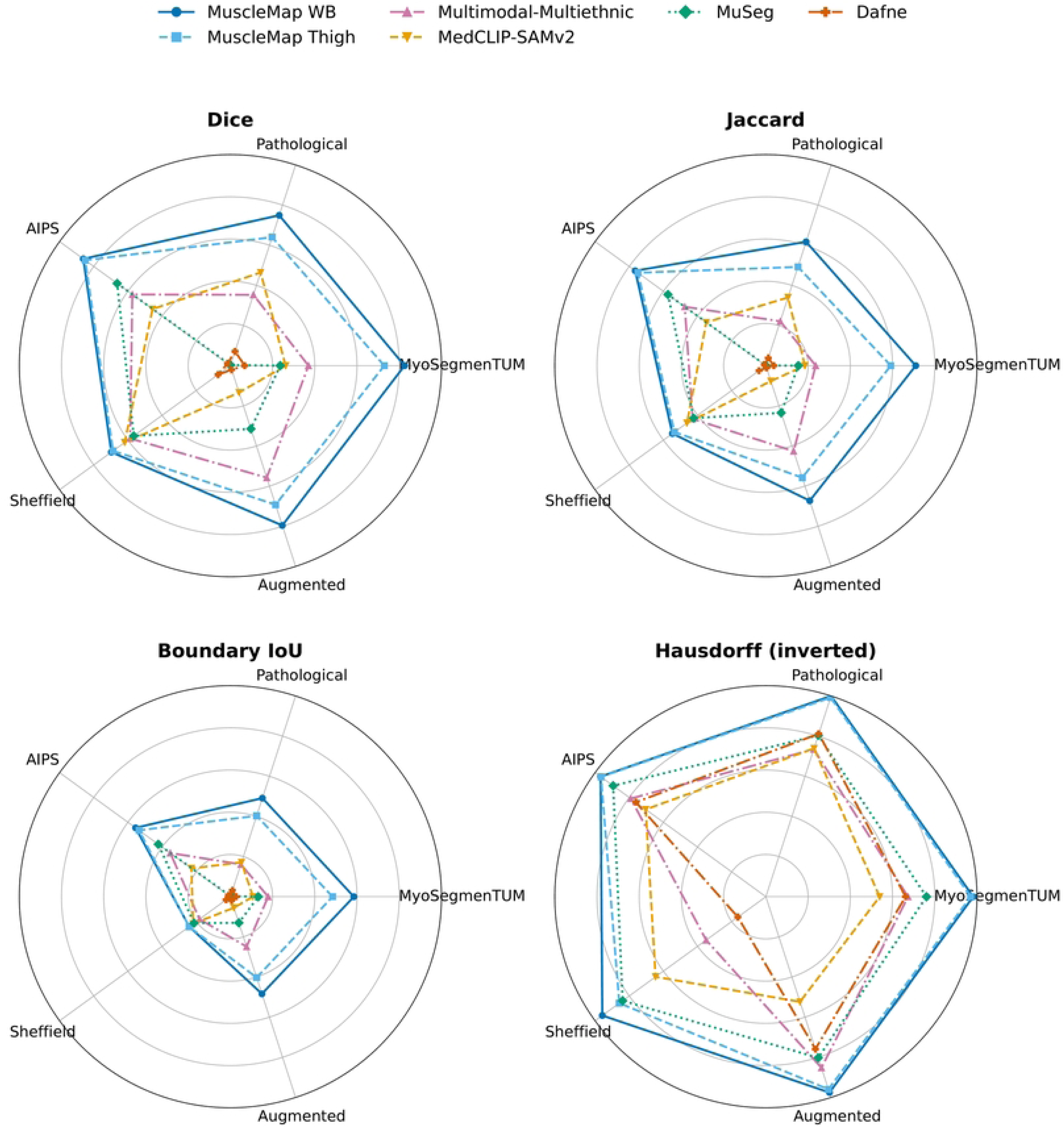
Results on datasets of various algorithms for main metrics. Dice, Jaccard and Boundary IoU are shown between their natural minimum and maximum values of 0 and 1, Hausdorff results are inverted and normalized to fit the consistent visual pattern wherein more area correlates with better results

**Fig 4.**
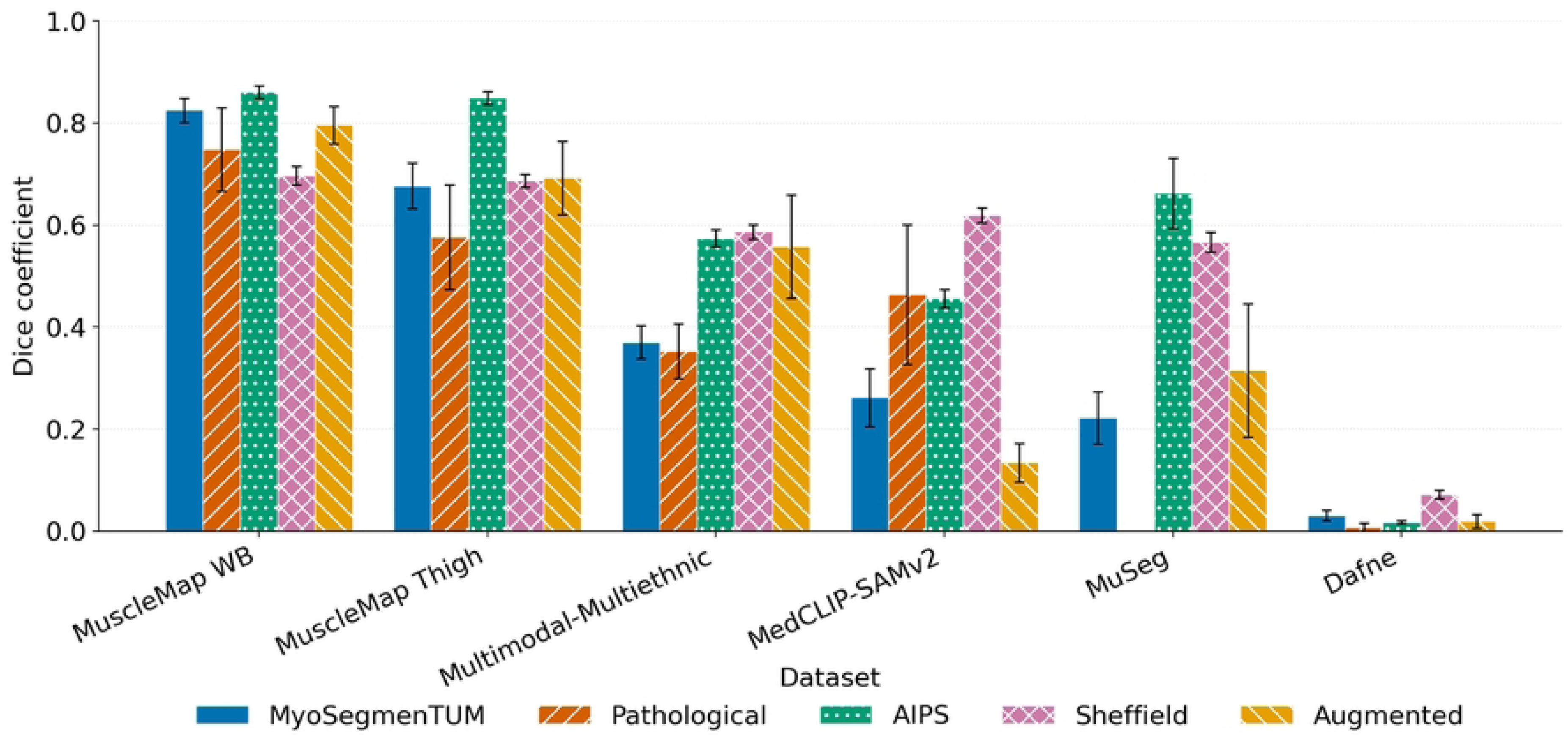
Results of various algorithms on various datasets on water sequence only are shown with black bars visualizing the 95% confidence intervals. MuSeg, which was designed to work on the whole Dixon sequence, completely mislabels the pathological subset on the water sequence, and therefore there is no bar.

**Fig 5.**
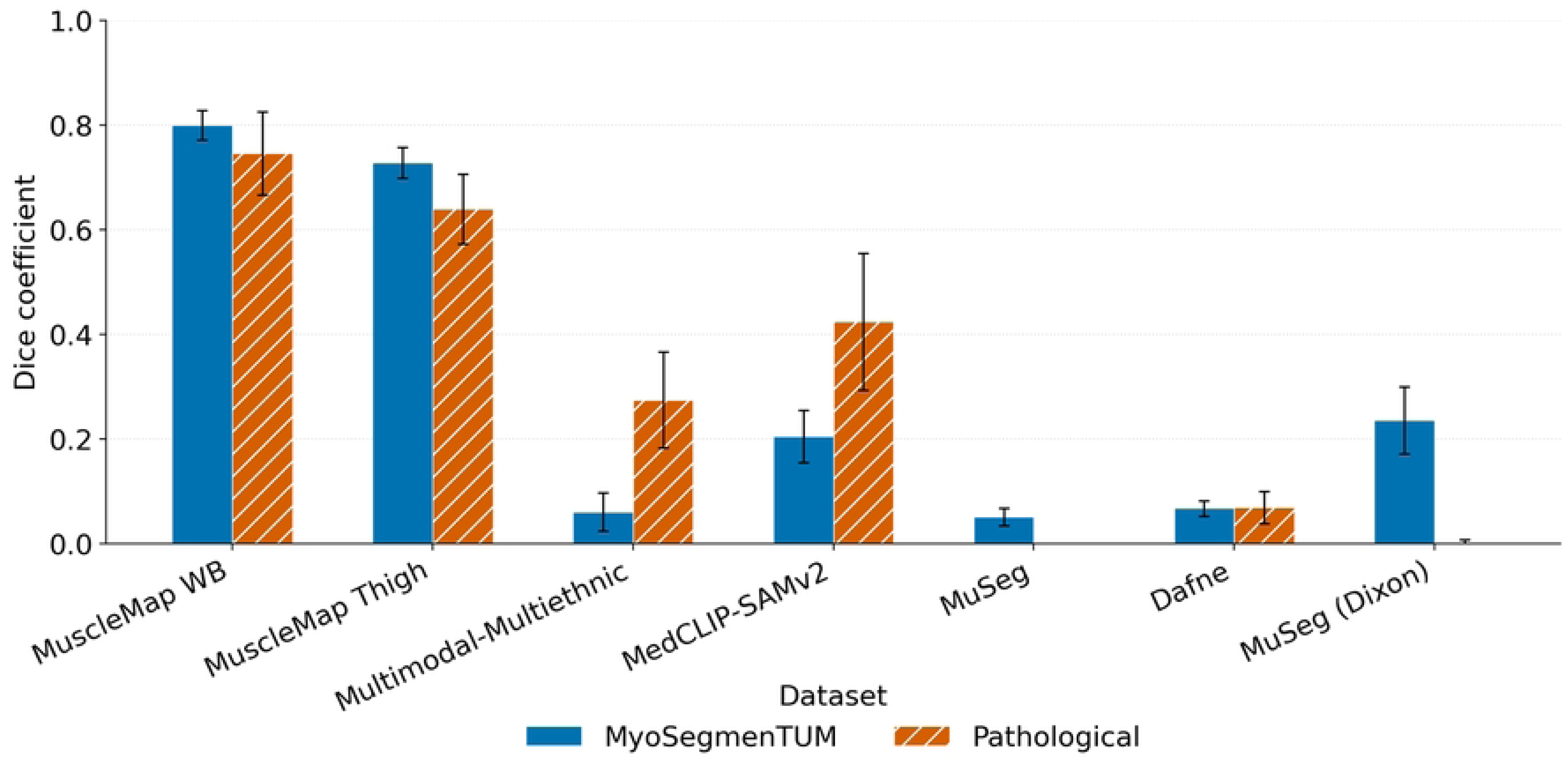
Results of various algorithms on various datasets are shown on fat fraction sequence only, black bars visualize 95% confidence interval. Note MuSeg, which was designed to work on the whole Dixon sequence, is also shown in terms of whole Dixon sequence although results are barely visible.

**Fig 6.**
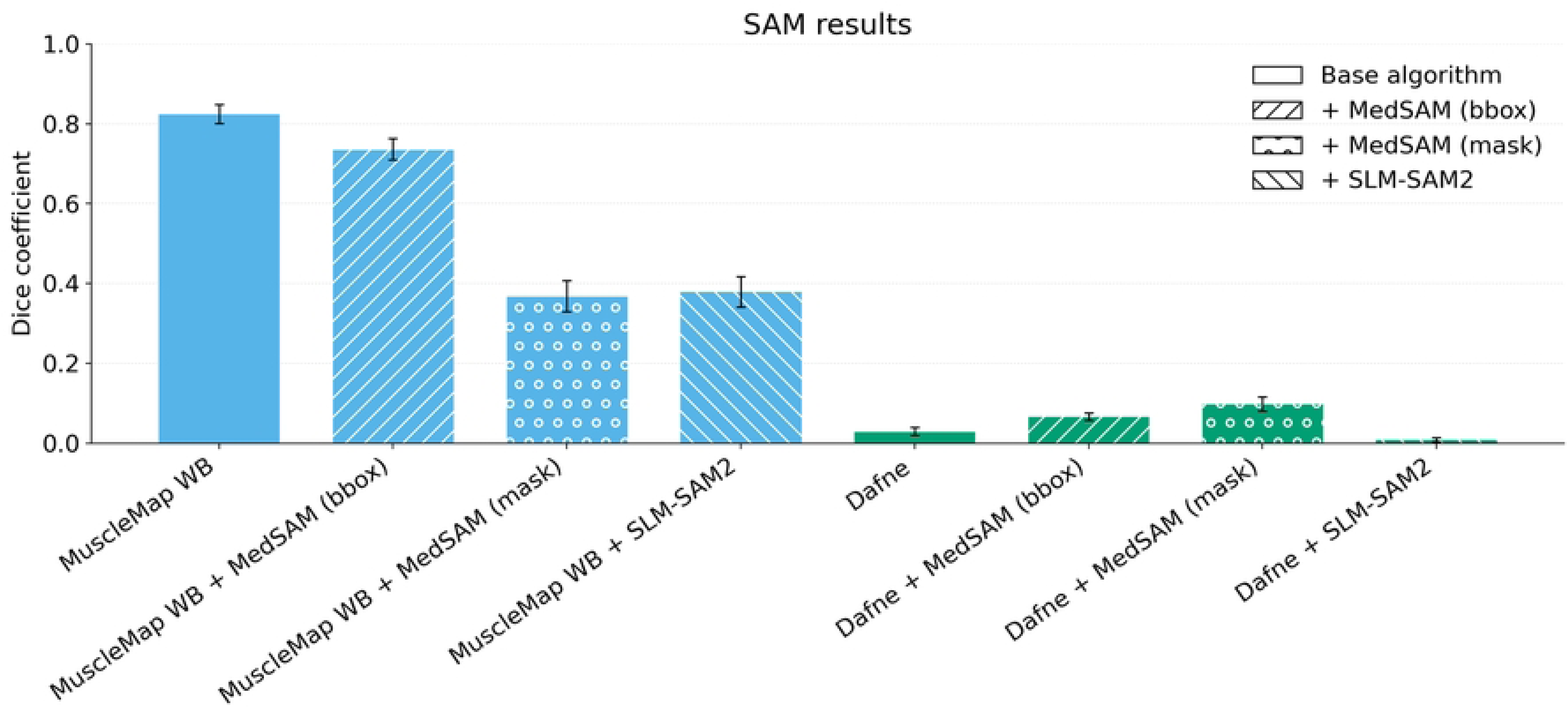
Results of various SAM algorithms applied to MuscleMAP WB or Dafne with MyoSegmenTUM water sequence only, black bars visualize 95% confidence interval. MedSAM was applied with bounding boxes, denoted by bbox or a mask.

Additional comparative parameters such as interslice Dice ratio, and binary cross entropy were examined. We also provide qualitative analysis.

We show the general Dice results in terms of algorithm results on water sequence, a specific image from the Dixon, as below:

Below we show general Dice results on fat fraction of the MyoSegmenTUM and Pathological subset. The graph also includes the whole Dixon sequence in the case of MuSeg.

Quantitative results are available in the supporting information (supplemental materials) [38].

Quantitative results from using SAM variants varied by implementation and algorithm applied to. A figure showing varying results of applying SAM variants to the best and worse performing algorithms shows the general trend of degrading results, but occasionally results improved in poor performing algorithms as below.

The qualitative results confirm the trend of MuscleMap outperforming other algorithms. Additionally, we noticed several qualitative patterns beyond those evident from overall metric values.

A key qualitative issue that emerged was the level of performance on pathological samples. The specific pathologies in the MyoSegmenTUM dataset, such as Limb Girdle Muscle Dystrophy, did not change all muscles equally. Thus results that average and include some relatively intact muscles can obscure truly poor performance. Because our ground truth labeling did not include all muscles, and grouped some e.g. the quadriceps, there was some inherent loss of this detail. However qualitative assessment showed that MuscleMap performed decently well on pathological muscles, and these often had questionable labeling of the ground truth. Unfortunately many other algorithms did not always perform as well on pathological samples. An example figure, of questionable ground truth labeling, good MuscleMap performance but poor performance from another algorithm, is shown in Figure 7.

**Fig 7.**
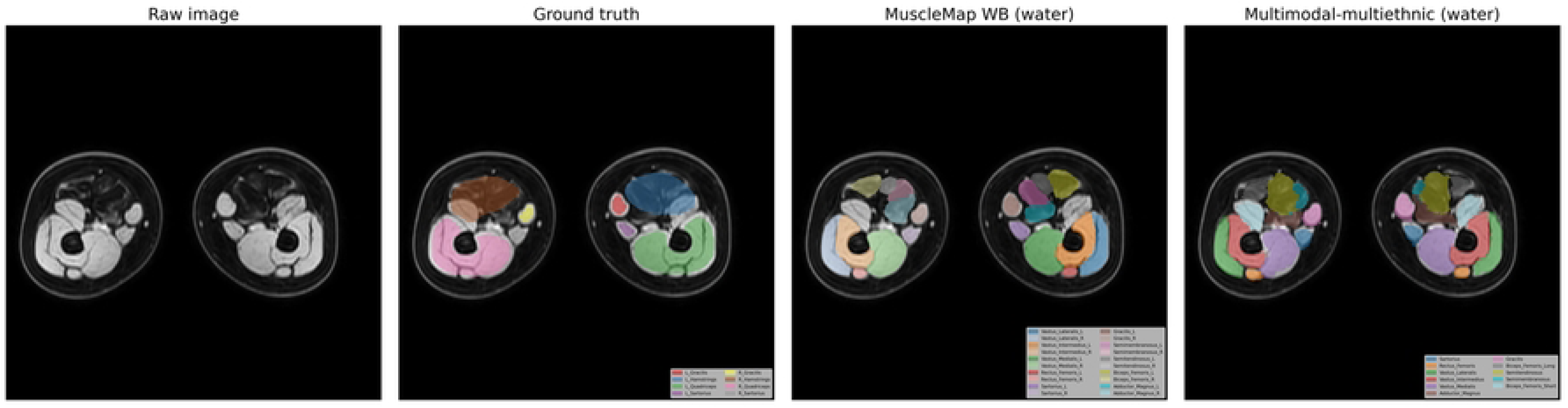
Labeled P001 1 WATER stack3 slice 23, notable for questionable ground truth labelling and good performance from MuscleMap on fat infiltrated muscle, poorer performance from multimodal-multiethnic

The better performance of Musclemap was a general qualitative trend but was not always the case. There is an example below where MuscleMap gives poorer performance on exceptionally small quadriceps muscles while Multimodal-Multiethnic shows better performance, as shown in Figure 8.

**Fig 8.**
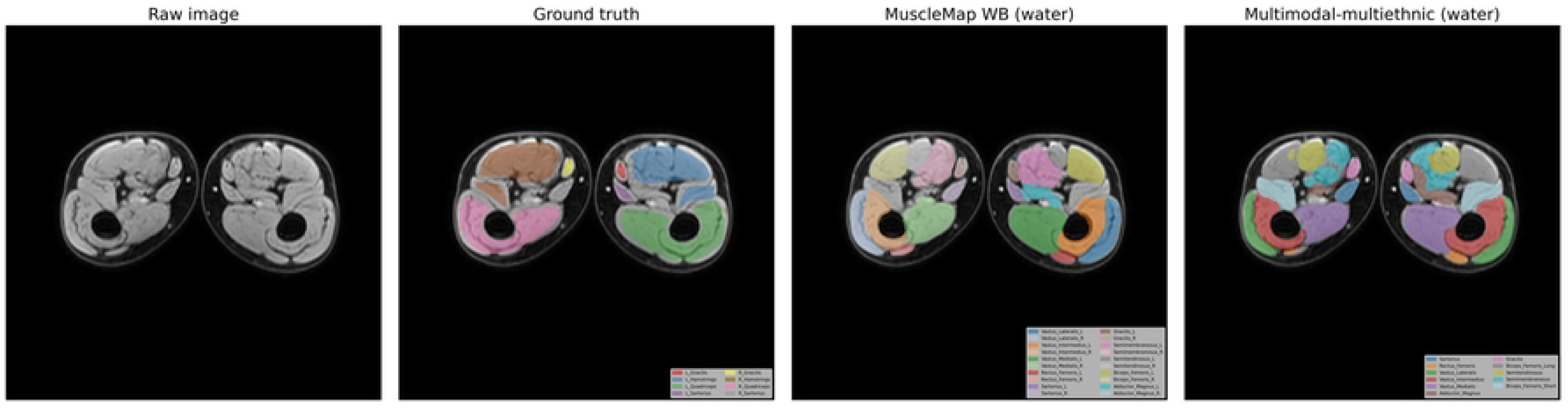
Labeled P003 1 WATER stack3 slice 21, notable for relatively small quadriceps and poorer performance from MuscleMap on these muscles, better performance from multimodal-multiethnic on the quadriceps

A second example of a pattern not captured by our quantitative metrics was that SAM variants would not only generally degrade performance, but specifically very often include bone in muscles even when cued with bounding boxes or points from well-performing algorithms. An example slice is shown in Figure 9.

**Fig 9.**
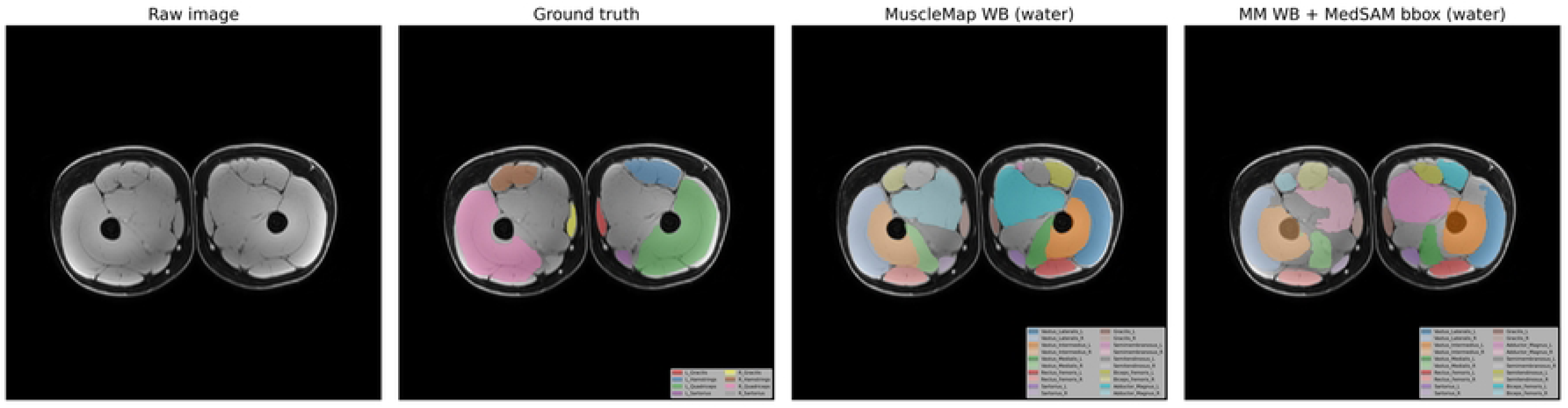
Labeled HV001 1 WATER stack1 slice 5

Some algorithms produced shapes illogical for a muscle on some slices, or in extreme cases lace-like or dotted label structures that would never logically be the formation of a muscle as muscles are continuous within themselves. Examples are shown in Figures 10 and 11.

**Fig 10.**
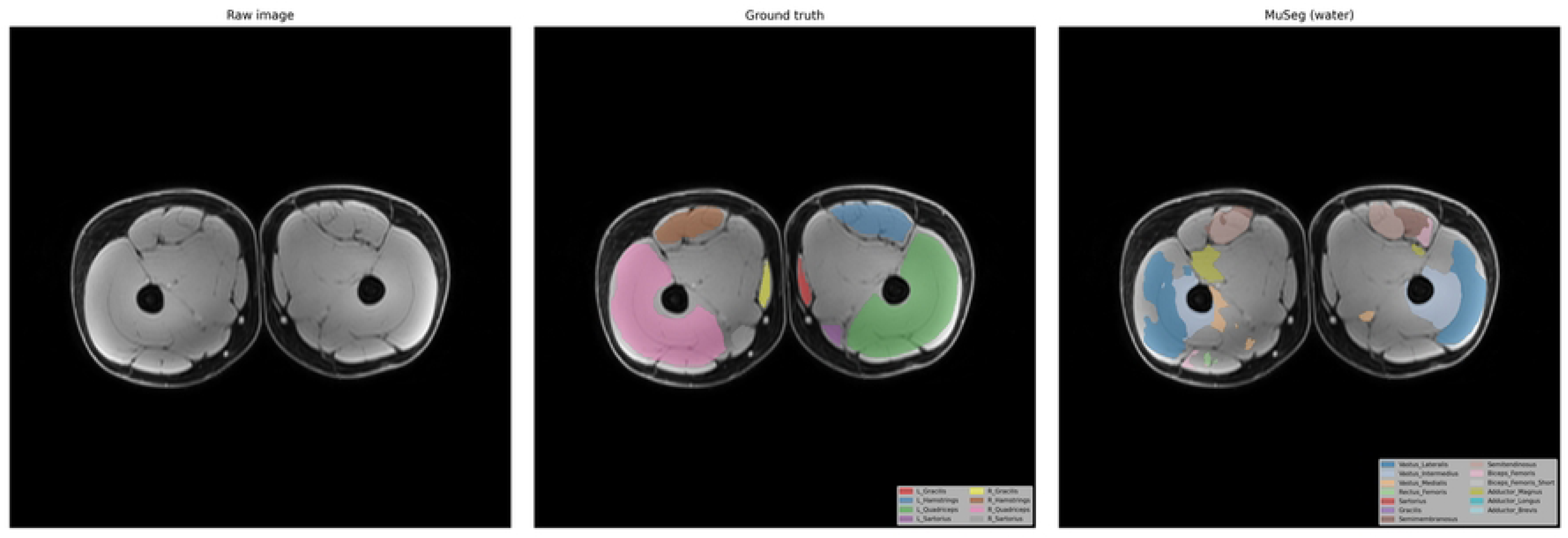
Labels with discontinuities in right leg from MuSeg as applied to HV001 1 WATER stack1 slice 2

**Fig 11.**
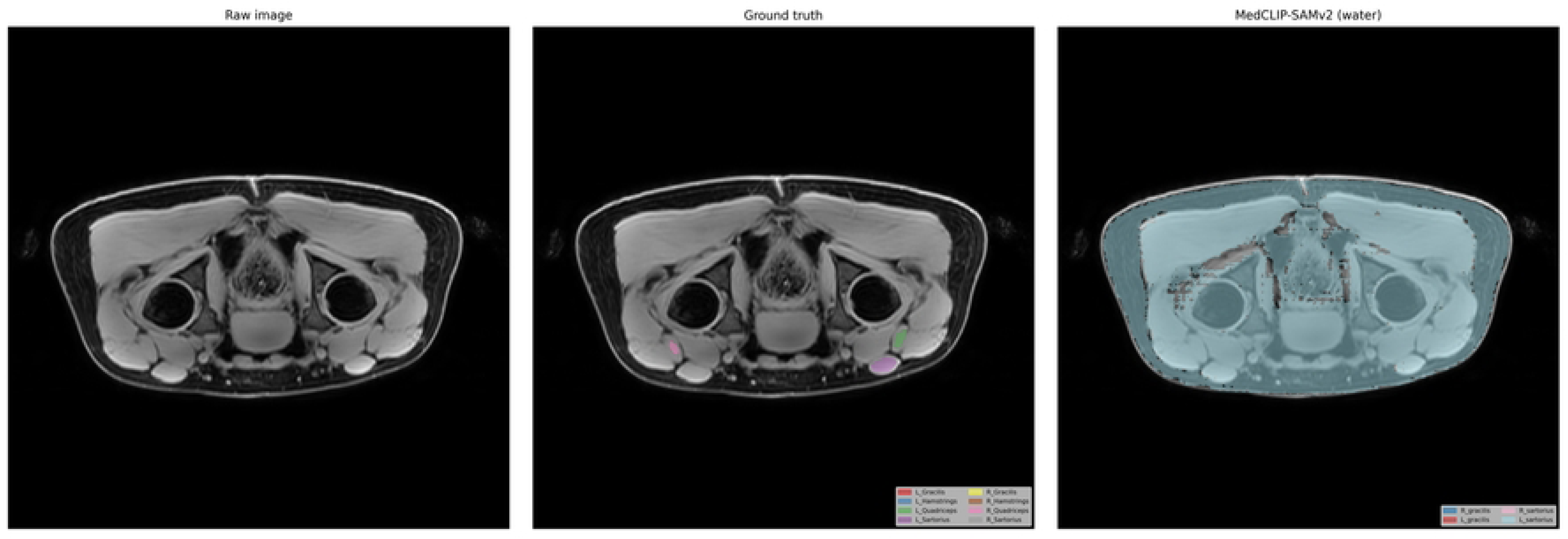
Example of label discontinuity from MedclipSAMv2 applied to HV001 1 WATER stack1 slice 48

These kinds of mistakes were almost never always the case, but rather would appear on some slices only. There was, however, with one exception in terms of an algorithm making continuous mistakes across every slice. The exception was the multimodal-multiethnic algorithm but only when applied to fat fractions. This combination always produced discontinuous, even speckled, labels outside of the muscle space. The failure of this algorithm in fat fraction is reflected in quantitative results as well.

It must be noted however that even discontinuous mistakes in segmentation can cause downstream problems such as extremely incorrect total muscle volumes, a feature of extreme clinical salience. Therefore, a desirable algorithm would use prior ‘knowledge’ of plausible muscle shapes to avoid this.

The question of whether adding an algorithm like SAM might correct such problems was addressed through our experiments. Unfortunately, the results of SAM variants were not always positive.

Qualitative results on all of MyoSegmenTUM revealed that all variants of SAM added to MuscleMap caused predictable changes that included inclusion of the femur into nearby muscles, and other labeling errors. Interestingly, this error of including the femur into muscles was also present in MedCLIP-SAMv2. This femur inclusion phenomenon suggests that something built in during training of SAM seems to somehow drive mislabeling.

Both the qualitative and quantitative results showed clearly that SAM variants did not enhance any of the best-performing algorithms. Thus on further testing SAM enhancement was not studied.

An ordinal table showing algorithm performances across datasets is shown in Table 4.

**Table 4.** Ordinal ranking of datasets per algorithm by Dice score (1 = best performance, 5 = worst). Algorithms only evaluated on a subset of datasets show fewer entries. Rankings based on best Dice score per algorithm per dataset.

| Algorithm | Rank 1 | Rank 2 | Rank 3 | Rank 4 | Rank 5 |
| --- | --- | --- | --- | --- | --- |
| MuscleMap WB | AIPS (0.861) | MyoSegmenTUM (0.825) | Augmented (0.796) | Pathological (0.749) | Sheffield (0.697) |
| MuscleMap Thigh | AIPS (0.850) | MyoSegmenTUM (0.729) | Augmented (0.693) | Sheffield (0.687) | Pathological (0.640) |
| Multimodal-Multiethnic Thigh Muscle MRI Analysis | Sheffield (0.587) | AIPS (0.574) | Augmented (0.558) | MyoSegmenTUM (0.370) | Pathological (0.353) |
| MuSeg | AIPS (0.663) | Sheffield (0.566) | Augmented (0.315) | MyoSegmenTUM (0.236) | Pathological (0.003) |
| MedCLIP-SAMv2 Text+Boxes | Sheffield (0.619) | Pathological (0.464) | AIPS (0.456) | MyoSegmenTUM (0.262) | Augmented (0.134) |
| Dafne | Sheffield (0.072) | Pathological (0.071) | MyoSegmenTUM (0.068) | Augmented (0.020) | AIPS (0.017) |

As Table 4 shows, most algorithms performed worst on the pathological dataset, although this was not universal. Despite performing slightly worse on pathological samples than on other datasets, MuscleMap and MuscleMap Thigh achieved much higher Dice scores on the pathological dataset than the other algorithms.

## Discussion

### Performance across methods and datasets

We evaluated all tools using multiple continuous metrics rather than a single composite score or binary threshold, as no universally accepted performance threshold exists for segmentation quality on any continuous metric.

Some tools performed poorly overall, while others were better but had notable patterns of performance such as working better in non-pathological samples or on certain muscles. Dafne performed consistently poorly as we implemented it, however Dafne is designed for interactive use: a user reviews and corrects each segmentation through the GUI, and those corrections feed back into the federated model. To evaluate all volumes under a single fixed model, we ran it via script without this correction step. Dafne’s poor performance therefore reflects its uncorrected output rather than what its intended workflow can achieve – a distinction that matters for scalability, since large studies cannot rely on per-image manual correction. It also exposes a risk: uncorrected predictions could in principle be contributed back to the federated server as if validated, degrading model quality over time. This concern extends to federated segmentation models generally – their training data cannot be fully cataloged, their most updated weights cannot be audited if updates are continuous, and continuous updates inherently compromise reproducibility.

Some algorithms performed worse on pathological samples and augmented data. Degraded performance on pathological cases is expected given their underrepresentation in training data. Degraded performance on augmented data is more concerning, as it could suggest reliance on similarity to training examples rather than learned anatomical features although we cannot rule out that specific augmentation choices contributed to performance drops. Visual inspection confirmed that augmented images remained anatomically plausible, but subtle artifacts from the augmentation pipeline may still have affected some tools disproportionately.

This pattern is also informative for a second reason: it is not uniform across tools, and that divergence is itself a finding the pathological-comparison alone did not reveal. MuscleMap (both variants) and MuSeg showed little to no drop in Dice between MyoSegmenTUM and the augmented set, while MedCLIP-SAMv2 lost roughly half its accuracy (as measured by Dice) under augmentation alone despite comparable or better performance on the Pathological Subset (Fig 3, Fig 4). Because the augmented images are anatomically plausible and derived from the same well-segmented subjects, this loss isolates positioning and orientation — not unfamiliar anatomy or disease presentation — as the variable driving the drop. Tools that degrade here behave as though they encode an implicit prior on patient position and framing learned from their training distribution, rather than a representation of muscle anatomy that is invariant to viewpoint.

These results are consistent with broader concerns about evaluation practices in medical image segmentation, where tools are often assessed on unrepresentative data using selectively favorable metrics [39–41]. Müller et al. [31] recommended visual inspection of segmentation outputs alongside quantitative metrics, and our experience supports this: qualitative review revealed systematic errors that standard metrics did not capture, such as SAM variants consistently incorporating bone into muscle labels.

We also observed that some tools performed well on certain muscles but poorly on others in a repeatable pattern, without obvious anatomical explanation. This warrants further investigation, to understand if there are universal predictors of poor performance in certain tools beyond the individual datasets already used, and underscores the value of per-muscle rather than aggregate-only reporting. We recommend that users of any segmentation tool visually compare outputs to the underlying images for at least a representative subset to identify such systematic trends.

### Practical usability

Domain-specific tools (MuscleMap, Dafne) were substantially easier to deploy, both offering graphical interfaces — MuscleMap via a 3D Slicer extension — that require no programming expertise. This is critical for adoption by clinical researchers. Another advantage of these tools is that they can be used to segment calf muscles as well, unlike the Multimodal-Multiethnic, or MuSeg tools, which means that researchers and clinicians who want to segment the entire lower limb to the ankle can use either of these tools without needing an additional tool for the calf.

All of the tools besides MuscleMap and Dafne presented significant usability barriers. SAM-based methods require conversion of 3D volumes to 2D slices, manual specification of bounding boxes or points for each muscle, and substantial anatomical knowledge. In practice, the manual effort required to prompt SAM-based tools may negate any time savings over manual segmentation. We partially automated this by generating prompts from MuscleMap predictions (leveraging its low false-positive rate), but this required software engineering as well as anatomical knowledge.

Most tools accept only specific MRI sequences and do not generalize across acquisition types. The Multimodal-Multiethnic tool, for example, segments well on fat or water images but produces nonsensical outputs on fat fraction images. Some tools further restrict usable inputs to specific echo times, severely limiting practical applicability. The Multimodal-Multiethnic tool also lacks laterality labels (left vs. right), though this is not relevant to all use cases. The MuSeg tool was designed to be used on the entire Dixon sequence, and performs better when used as designed, however it does not perform well on the MyoSegmenTUM dataset.

All evaluated tools produce binary masks rather than probabilistic outputs. Probabilistic segmentation — where each voxel receives a confidence estimate — has been explored in brain MRI [42, 43] and in general computer vision via the Probabilistic U-Net [44], but has not been adopted for musculoskeletal imaging. Such an approach could allow users to apply task-appropriate confidence thresholds and merits future investigation.

### Limitations

Our usability assessment is only qualitative; a formal usability study with structured user evaluation would strengthen these observations.

Several methodological limitations affect our quantitative comparisons. None of the three source datasets report formal measures of inter- or intra-rater annotation reliability in terms of ground truth which was produced by single expert annotators (with radiologist supervision, where reported) rather than through consensus or repeated-annotation protocols. We noted some limitations in the Sheffield dataset labeling specifically. This is a limitation when interpreting all reported accuracy metrics as reflecting deviation from the ground truth metric. We did not evaluate every muscle in every dataset; some algorithms performed markedly worse on specific muscles, and a complete per-muscle analysis across all datasets would provide a more detailed picture. The datasets themselves are geographically limited — no data from Africa or the Americas is included — meaning that potential biases related to ethnicity or body composition remain unexplored. Relatedly, known sex differences in muscle composition (e.g., higher fat infiltration in female muscles [45]) may interact with tool performance, but we did not stratify results by sex because differences in muscle cross-sectional area would confound such comparisons without careful normalization.

Comparing tools that expect different input modalities (fat fraction, water, Dixon) introduces inherent asymmetry. Tools using multi-channel inputs can exploit inter-channel contrast that single-channel tools cannot. We report results on each tool’s intended input type, which reflects realistic use but makes direct algorithmic comparison imperfect. A fully controlled comparison would require retraining all tools on identical inputs, which was beyond the scope of this study. We did not evaluate inference speed, computational cost, environmental impact, or susceptibility to automation bias — all relevant considerations for clinical deployment.

Another limitation of our work is that we report performance on pathological samples as a whole; however, these samples do not represent cases in which every muscle is necessarily affected. Because of the potential of unaffected muscles, overall performance could be misleading. We cannot conclusively judge which muscles had pathological changes because we did not have clinical information in the datasets to confirm which muscles were truly affected, and effects might only be visible at the radiomic level, thus not categorizable for a valid data analysis of only affected muscles.

Finally, our evaluation captures a snapshot of a rapidly evolving landscape. Tools may have been updated since testing, and new tools may have become available. The evaluation framework and datasets used here are publicly available [29] to facilitate periodic re-evaluation as the field progresses.

## Conclusion

We present an independent benchmark of eight open-source segmentation tools for thigh muscle MRI, evaluated across four datasets encompassing healthy subjects, pathological cases, and augmented out-of-distribution data. Performance varied substantially across tools, datasets, and individual muscles.

Domain-specific U-Net models trained on large, multi-site datasets — particularly MuscleMap — consistently outperformed foundation models and newer general-purpose architectures. Two specific findings stand out. First, the thigh-specific MuscleMap model unexpectedly underperformed its whole-body counterpart, suggesting that broader training data can compensate for reduced task specificity. Second, augmenting existing tools with SAM variants (MedSAM, slmSAM) often degraded rather than improved segmentation quality across many tested configurations, despite SAM’s established strengths.

Most tools did not show dramatically reduced performance on pathological cases and augmented data. The opposite would have suggested that current models heavily depended on similarity to training examples rather than generalizable anatomical features, though we note this would have been a plausible explanation requiring further investigation. Nonetheless the best models, MuscleMap models, do technically have slightly lower performance metrics on pathological data although the difference is within confidence intervals. This closeness of numbers may reflect that in diseased patients not every muscle is affected, thus the relatively healthy muscles that segment well can obfuscate the quantitative trends. If this trend towards worse performance on pathological data is confirmed on other datasets, it would raise significant concerns about deploying these tools on underrepresented pathologies or populations not well reflected in existing training sets.

While technically no single tool was optimal across all metrics and use cases, we can look at primary metrics as indicative of performance, as secondary metrics can be misleading due to artifacts e.g. a zero false-positive rate can represent almost no segmentation. Upon looking at primary metrics, we conclude that for applications prioritizing broad muscle labeling with acceptable accuracy, MuscleMap is the strongest current option. However, even MuscleMap’s outputs are less precise than hand labeling, particularly at muscle boundaries; for applications requiring exact delineation – such as radiotherapy planning, where precise structure borders directly affect treatment fields – no evaluated tool is yet reliable enough for unsupervised use. We therefore recommend that any automated segmentation be paired with visual inspection and, where boundary precision is clinically critical, manual review and correction.

These findings point to a clear need for segmentation approaches that can generalize from limited training data to novel populations and pathologies. Our evaluation framework and datasets are publicly available [29] to support ongoing benchmarking as the field evolves.

## Supporting information

Supporting data can be found on Zenodo [38]. Additionally, some supporting figures are available with this publication.

**S1 Fig. Results on the MyoSegmenTUM dataset and its pathological subset for various algorithms.** Dice scores by sequence (water, fat fraction) for each segmentation algorithm.

**S2 Fig. Results on the AIPS and Sheffield datasets for various algorithms.** Dice scores by sequence for each segmentation algorithm.

**S3 Fig. Results on the augmented dataset for various algorithms.** Dice scores by sequence for each segmentation algorithm.

**S4 Fig. Example of bone inclusion in a SAM-variant segmentation.** Labeled slice 5 of HV001 1 WATER stack1.

**S5 Fig. Example of label discontinuity from MuSeg.** Labels with discontinuities in the right leg, slice 2 of HV001 1 WATER stack1.

**S6 Fig. Example of label discontinuity from MedCLIP-SAMv2.** Slice 48 of HV001 1 WATER stack1.

## Data Availability

Related data are available at 10.5281/zenodo. 21840979 (on Zenodo). Code is open and also archived on Zenodo at https://zenodo.org/records/20787771

https://doi.org/10.15131/shef.data.20440164

https://zenodo.org/records/21840980

https://doi.org/10.6084/m9.figshare.31042489

https://osf.io/svwa7/?view_only=c2c980c17b3a40fca35d088a3cdd83e2

## Acknowledgments

This research was supported in part by Lambda, Inc. Lambda, Inc. had no role in the design of the study, analysis, interpretation of data, or writing of the manuscript for publication.

## Notes

### Competing Interest Statement

The authors have declared no competing interest.

### Author Declarations

As the work was based on open previously anonymized images, there was no IRB approval needed.

